# Pembrolizumab, Temozolomide and HSPPC-96 Vaccine in Newly Diagnosed Glioblastoma Post-Chemoradiation: Results from a Multi-institutional, Phase 2, Randomized, Placebo-Controlled Trial

**DOI:** 10.64898/2026.06.22.26354817

**Authors:** Byram H. Ozer, Scott M. Lindhorst, Ryan T. Merrell, Christopher R. Trevino, Jeremy D. Rudnick, Nicolas G. Avgeropoulos, Naren Ramakrishna, Simon Khagi, Yasmeen Rauf, Tobias Walbert, Edward Pan, Michael Youssef, Karen L. Fink, Jacob J. Mandel, Lynne P. Taylor, Howard Colman, Erin M. Dunbar, Nina Paleologos, Eric C. Burton, Jing Wu, Heather E. Leeper, Javier Gonzalez, Marta Penas-Prado, Jeffrey J. Raizer, Eleonora Veglia, Sandra Craig, Ying Yuan, Claudia Chambers, Kathleen Wall, Ewa Grajkowska, Tito Mendoza, Terri S. Armstrong, Mark R. Gilbert

**Affiliations:** Neuro-Oncology Branch, National Cancer Institute/National Institutes of Health, Bethesda, Maryland, USA; Department of Neurosurgery, Medical University of South Carolina, Charleston, South Carolina, USA; Department of Neurology, NorthShore University Health System, Chicago, Illinois, USA; Department of Neurology, Cedars-Sinai Medical Center, Los Angeles, California, USA; Department of Hematology and Medical Oncology, Orlando Regional Medical Center, Orlando, Florida, USA; Department of Radiation Oncology, Orlando Regional Medical Center, Orlando, Florida, USA; Department of Neurology and Neurosurgery, University of North Carolina at Chapel Hill, Chapel Hill, North Carolina, USA; Department of Neurology and Neurosurgery, Henry Ford Health System, Detroit, Michigan, USA; Department of Neurology, University of Texas Southwestern, Dallas, Texas, USA; Department of Neurology/Neuro-Oncology, Baylor Scott&White, Baylor University Medical Center, Houston, Texas, USA; Department of Neurology, Baylor College of Medicine, Houston, Texas, USA; Department of Neurology, Alvord Brain Tumor Center, University of Washington and Fred Hutchinson Cancer Center, Seattle, Washington, USA; Department of Neurosurgery, Huntsman Cancer Institute, Salt Lake City, Utah, USA; Piedmont Brain Tumor Center, Atlanta, Georgia, USA; Division of Neurology, Advocate Medical Group, Downers Grove, Illinois, USA; Department of Neurology, Feinberg School of Medicine, Northwestern University, Chicago, Illinois; Department of Biostatistics, MD Anderson Cancer Center, Houston, Texas, USA; Scientific Leadership and Research Manager, MSD/Merck&Co, Inc., Rahway, New Jersey, USA; Director of Manufacturing and Logistics, Agenus, Lexington, Massachusetts, USA

**Keywords:** Glioblastoma, HSPPC-96, Pembrolizumab, Clinical Trial, Cancer Vaccine

## Abstract

**Background:** GBM is one of the most common and most aggressive brain tumors in adults, and upfront standard of care treatment has limited efficacy. Immune checkpoint inhibitor strategies have significantly improved outcomes in various solid tumors but have not proven effective in GBM, suggesting other strategies may be needed to realize their full potential.

**Methods:** GBM patients were treated with upfront standard of care chemoradiation with temozolomide and pembrolizumab, followed by adjuvant temozolomide and pembrolizumab for six nine-week cycles. Depending on production of sufficient vaccine, patients were randomized into HSPPC-96 vaccine or placebo group (q4 weeks) while those with failed vaccine production continued on study unblinded as an ancillary group. The primary objective was overall survival at one year, and secondary endpoints were progression-free survival at six months, overall and progression-free survival, radiographic response, and tolerability by patient-reported outcomes and adverse event documentation.

**Results:** 90 patients were screened, 32 were treated (8 vaccine, 9 placebo, 15 ancillary), and 26 were evaluable for radiographic responses prior to accrual termination. The study did not meet its primary endpoint of overall survival at one year (65.5% in vaccine group, 75% in placebo). Progression-free endpoints were mildly improved in the vaccine group but were not significant, and response rates were not significantly different. The regimen was well-tolerated and safe.

**Conclusions:** Though limited by early discontinuation, these findings do not support the combination of pembrolizumab and HSPPC-96 vaccine with standard of care therapy.

**Trials Registration:** ClinicalTrials.gov identifier: NCT03018288

**Key Points:**

- Newly diagnosed glioblastoma (GBM) has one standard of care treatment option with known limitations.
- Combination of standard of care therapy with immunotherapeutics consisting of pembrolizumab and the personalized cancer-derived HSPPC-96 vaccine is safe and well-tolerated in newly diagnosed GBM
- This regimen demonstrated no survival benefit, but the vaccine safety profile recommends consideration of alternative combinatorial strategies

**Importance of the Study:** In this phase 2, multicenter, placebo-controlled, double-blinded trial of newly diagnosed GBM patients, the addition of pembrolizumab and patient tumor-derived HSPPC-96 vaccine to standard of care chemoradiation and adjuvant temozolomide was not effective even accounting for early termination due to accrual issues during the Covid pandemic. However, with good safety profile and tolerability, HSPPC-96 vaccine could be considered in combination with other immunotherapeutic strategies in future GBM trials.

## Introduction

Glioblastoma IDH WT, WHO CNS Grade 4 (GBM) is the most common primary brain malignancy in adults^1^. This is a uniformly aggressive tumor with a median survival of 12-15 months with standard of care treatment. GBMs without O6-methylguanine-DNA methyltransferase (MGMT) promoter methylation are resistant to standard of care temozolomide and tend to progress rapidly and lead to early mortality^2^. Unfortunately, since the establishment of temozolomide combination with radiation therapy in 2005^3^, there has been little progress in the upfront management of GBMs, highlighting a need for therapeutic improvements for this rare and deadly disease.

Immunotherapy has revolutionized treatments in many solid tumors, including melanoma, lung cancer, renal, and colorectal cancers^4^. Immune checkpoint inhibitors are a class of antibodies that target the interaction between leukocyte expression of programmed death-1 (PD1) and aberrant expression of programmed death ligand 1 (PDL1) on tumor cells, whose interaction results in the suppression of activated T-lymphocytes normally involved in immune surveillance and neoplastic cell clearance^5^. Pembrolizumab is one such immune checkpoint inhibitor developed for use in solid tumors both as a combination and monotherapy. Although there is evidence of improved clinical outcomes in solid tumors with brain metastases^6,7^, its efficacy as a monotherapy has not been recapitulated in GBM, likely due to a combination of low tumor mutation burden, minimal surface PDL1 expression, and relatively inactive or “cold” anti-tumor immune microenvironments^8^. Strategies to boost the immunogenicity of GBM or its microenvironment are postulated to be a necessary precondition for unlocking the full potential of immune checkpoint inhibitor therapy in this disease.

Heat shock proteins (HSPs) are molecules that respond to cellular stress, counteract abnormal protein folding, and modulate immune responses by functioning as intracellular chaperones that deliver tumor antigens to antigen presenting cells (APCs) for immune activation. The Heat Shock Protein-Peptide Complex-96 (HSPPC-96) vaccine is an autologous tumor-derived vaccine composed of the 96 kDa heat shock protein glycoprotein-96 (HSP gp-96) in complex with autologous tumor-derived peptides^9^. The gp96 component is a highly conserved, abundantly expressed, nonpolymorphic stress protein that is found in every body tissue, but isolated from tumor is expected to be in complex with a diverse repertoire of peptides with opportunities to present peptides unique to the tumor cell of origin. The injected vaccine interacts with APCs and, once internalized, is chaperoned to major histocompatibility complex (MHC) class I and II molecules in intracellular compartments to be expressed at the cell surface^10^. These activated APCs in turn travel to lymph nodes for presentation to naïve T-cells. This leads to widespread immunological activation, including CD8+ and CD4+ positive T-cell responses against the presented antigen, cytokine and chemokine release by dendritic cells and macrophages, maturation of dendritic cells, and activation of natural killer cells. HSPPC-96 vaccines are therefore an attractive strategy for immunological priming as a cancer-specific vaccine^11,12^.

In this multi-institutional, randomized, double-blinded, placebo-controlled, Phase 2 trial, we treated MGMT unmethylated GBM with standard chemoradiation regimen (radiation + temozolomide) together with pembrolizumab and subsequently compared the effect of adjuvant temozolomide and pembrolizumab, with and without the addition of an autologous HSPPC-96 vaccine created from patient autologous tumor tissue. Our goal was to evaluate the clinical responses and toxicities of the immunological combination therapy while providing standard of care management.

## Methods

### Patient Accrual

This was a multi-institutional trial run through the Brain Tumor Trials Collaborative (BTTC) with the National Cancer Institute serving as the lead institution and coordinating center. The study was randomized, double-blinded, placebo-controlled and enrolled adult patients with suspected GBM. Because vaccine production required at least 7 grams of tumor tissue and processing to maintain tissue integrity for transport to vaccine-preparation facilities, a two-step accrual process was put into place. The pre-surgical process required registration both at the local treatment and central coordinating (NCI) centers based on strong radiological suspicion of GBM, preliminary assessment of feasibility to remove >80% of tumor tissue and expectation of adequate volume for vaccine manufacture, and adequate performance status (Karnofsky Performance Status (KPS) ≥70%). The post-surgical registration process required histopathological confirmation of GBM IDH WT MGMT promoter unmethylated, appropriate post-craniotomy recovery, organ, and bone marrow function, and plans to initiate standard of care chemoradiation treatment within 2-6 weeks post-surgery. Standard exclusion criteria regarding previous cancers, brain tumor, or immunotherapeutic treatments also applied. After this second registration step, eligible patients would start standard of care chemoradiation with pembrolizumab added to the regimen. Vaccine was manufactured in parallel using the resected tumor tissue at Agenus facilities.

Approximately one month after completion of chemoradiation with pembrolizumab, patients who had successful vaccine production during the initial chemoradiation period, with sufficient vaccine defined as a minimum of four injections, were subsequently randomized into two groups: those who received vaccine injections (Vaccine Group) and those who received placebo injections (Placebo Group). This randomization occurred as a third registration step in direct coordination between the central registration site (NCI) and the manufacturer (Agenus), who would subsequently send blinded vaccine or placebo directly to the treatment site. Accrued patients where vaccine production was unsuccessful were maintained on pembrolizumab treatment through the adjuvant chemotherapy treatment period in order to maintain study participation and follow-up but had no corresponding vaccine or placebo injection and were not blinded to their treatment (Ancillary Group).

### Investigational Agent Administration

Enrolled patients started therapy after adequate post-surgical recovery. On day one, temozolomide (TMZ) was started orally at 75 mg/m^2^ every day in conjunction with week-daily radiation with a goal total dose of 60 Gy (33 fractions at 180 cGy or 30 fractions at 200 cGy). To this standardized regimen, pembrolizumab (Pembro, provided by MSD/Merck) was infused intravenously (IV) at 200 mg starting the first treatment day and continuing every three weeks (e.g., Days 22 and 43).

For those patients for whom sufficient vaccine was produced (provided by Agenus), HSPPC-96 vaccine or placebo was administered by a single intradermal injection, and this continued every four weeks (timed 21 days after TMZ cycle initiation) for 6 cycles or until the vaccine supply was depleted unless progression occurred first.

Irrespective of vaccine availability/administration, all enrolled patients started their adjuvant regimen at four weeks post-radiation. The standard adjuvant regimen was modified to a nine-week regimen consisting of temozolomide administered on Days 1-5 at 150 mg/m^2^, followed by 200 mg/m^2^ on Days 29-33 and all subsequent cycles as tolerated. To this was added pembrolizumab 200 mg IV administered days 1, 22, and 43 in the nine-week cycle. Once six cycles (i.e., 54 total weeks) were completed, patients could continue pembrolizumab alone for an additional 12 months. In rare cases where vaccine product was still available, continuation of vaccinations every four weeks was permitted.

### Data Collection

Baseline assessments (post-surgery and pre-treatment) included physical and neurological exam with KPS, standard laboratory evaluations including thyroid evaluation, as well as the M.D. Anderson Symptom Inventory-Brain Tumor (MDASI-BT) patient-reported outcome (PRO) measures. While on trial, physical, neurological, and KPS evaluations were performed every 3 weeks during chemoradiation, then every four weeks thereafter. MRIs were obtained pre- and post-surgery, once during the post-chemoradiation recovery period, and then every 9 weeks in conjunction with the beginning of each nine-week cycle. Hematological labs were performed weekly and organ function, including thyroid, every three weeks during active treatment. MDASI-BT was administered at baseline, post-chemoradiation prior to adjuvant treatment administration, and subsequently at every imaging visit. CTCAE version 5.0 was used for adverse event reporting and was collected at every in-person visit.

As part of the exploratory objectives, efforts were made to collect blood for immunophenotyping at baseline (pre-chemoradiation), post-radiation, as well as on Day One of each even cycle through eight cycles. When collected, samples were sent to the central processing center (NCI) for storage and analysis at the completion of the study. Any relevant results will be reported separately.

### Statistical Methods

Overall survival (OS) at 12 months (OS-12) was the primary objective, and secondary objectives included progression-free survival (PFS) at 6 months (PFS-6) and OS. Additional secondary objectives included safety and tolerability, including patient-reported outcomes, and radiographic response rates. Kaplan-Meier method is used for the analyses of progression free survival (PFS) and overall survival (OS). Median PFS and median OS with their 95% confidence interval (CI) are provided for each cohort (HSPPC-96 vaccine/placebo/ancillary). Rates of PFS at 6 month and rates of OS at 6 months, and 2 years, along with their 95% CI are provided for each cohort. Chi-square was used to compare the demographic variables, and Chi-square Log Rank testing was used to compare OS and PFS between two experimental arms (HSPPC-96 vaccine/placebo). The Fisher Exact test is used to compare the one-year OS between two experimental arms. Rates of best response to treatment are summarized by category and 95% CI are provided for each cohort. MDASI-BT patient-reported outcomes were analyzed using linear mixed models fitted using autoregressive covariance structure, which assumes relationships among symptoms decrease progressively over time. Pairwise comparisons among treatment arms and among time points were subsequently analyzed using the Bonferroni method.

### Ethics Statement

The trial was carried out in accordance with International Council for Harmonisation Good Clinical Practice (ICH GCP) and all investigators and site staff responsible for the conduct, management, or oversight of this trial completed Human Subjects Protection and ICH GCP Training. The protocol, consent, and related materials were all reviewed and approved by the NIH Clinical Center IRB (17C0034).

## Results

### Patient Population

90 total patients were screened for the trial with 32 patients incorporated into one of three groups: eight patients were randomized into the HSPPC-96 vaccine group and nine patients into the placebo group, while 15 patients with insufficient vaccine production were assigned into the pembrolizumab-only group (ancillary group) and included for assessment of primary and secondary endpoints. Importantly, accrual was halted after the final patient was registered on 3/3/2021 because of logistical issues related to the ongoing pandemic. Of the 32 patients, all patients were available for survival analyses while twenty-six patients were evaluable for response rates (Figure 1). Patient demographics (Table 1) show that most enrollees were over the age of 50 and overwhelmingly male with roughly similar median age of entry and with good performance status. Though there were individual differences in tumor location between the experimental cohorts (majority right-sided and parietal in the vaccine group, majority right-sided and frontal in the placebo group, and left-sided frontal and temporal in the ancillary group), as well as differences in extent of resection (more gross-total resections in placebo compared to vaccine and ancillary groups) that is known to affect clinical outcomes in GBM^13^, these differences did not meet criteria for statistical significance.

**Figure 1:**
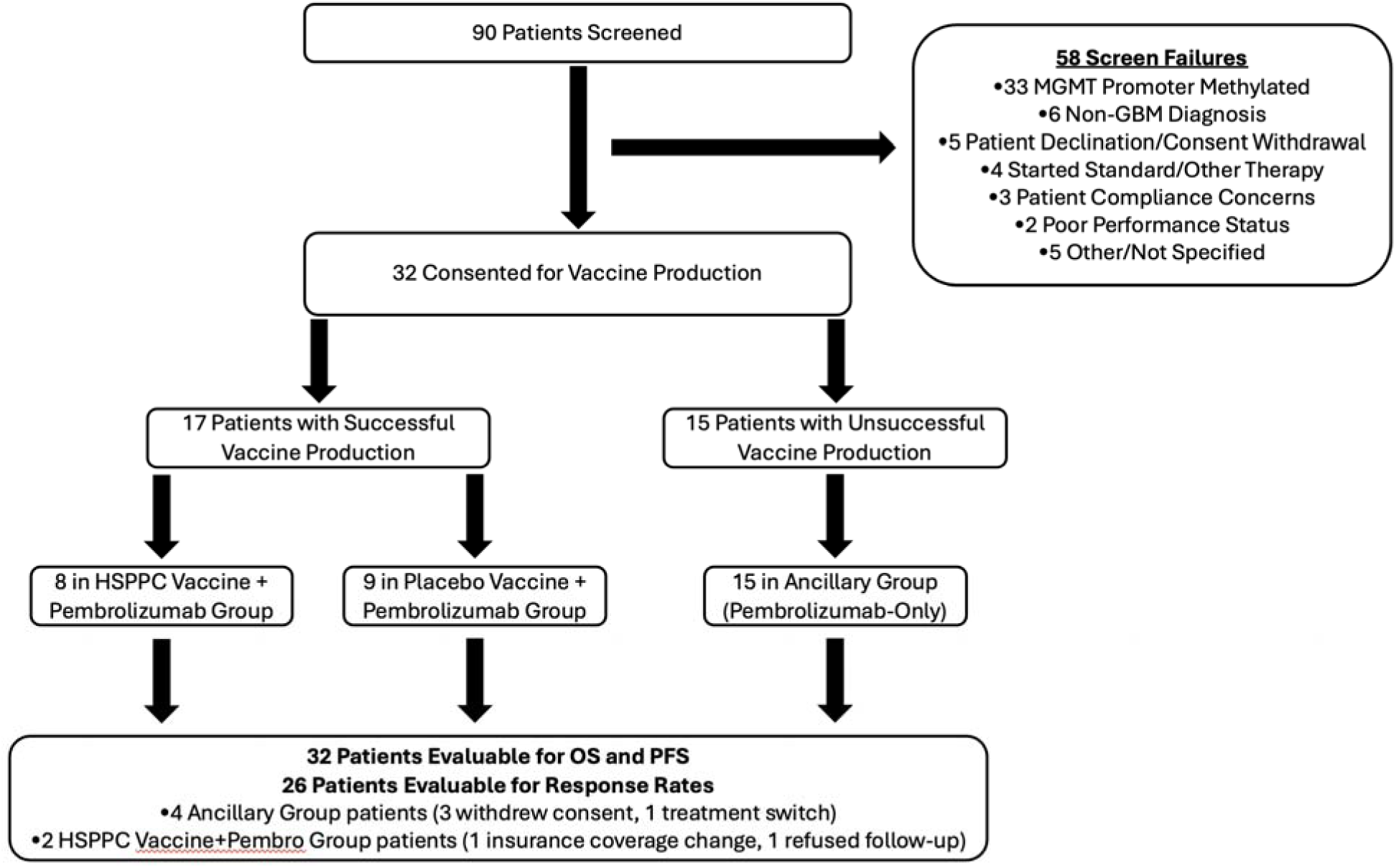
Schema outlining patient accrual and randomization process as well as sources of screen failure and characteristics of patients removed from response rate analyses.\

**Table 1.** Participant Demographics.

|  | HSPPC-96 Vaccine |  | Placebo |  | Ancillary |  | Total |
| --- | --- | --- | --- | --- | --- | --- | --- |
| Age | n | % | n | % | n | % | n |
| <50 | 1 | 12.5% | 4 | 44.4% | 2 | 13.3% | 7 |
| ≥50 | 7 | 87.5% | 5 | 55.6% | 13 | 86.7% | 25 |
| Mean Age at Entry (SD) | 64.0 | (11.9) | 58.4 | (12.6) | 58.7 | (8.8) |  |
| Sex |  |  |  |  |  |  |  |
| Male | 7 | 87.5% | 7 | 77.8% | 9 | 60.0% | 23 |
| Female | 1 | 12.5% | 2 | 22.2% | 6 | 40.0% | 9 |
| Race |  |  |  |  |  |  |  |
| Asian | 0 | 0.0% | 1 | 11.1% | 2 | 13.3% | 3 |
| Black/African American | 0 | 0.0% | 1 | 11.1% | 1 | 6.7% | 2 |
| White | 8 | 100.0% | 7 | 77.8% | 12 | 80.0% | 27 |
| KPS at Baseline |  |  |  |  |  |  |  |
| 90-100 | 5 | 62.5% | 7 | 77.8% | 8 | 53.3% | 20 |
| 70-80 | 3 | 37.5% | 2 | 22.2% | 6 | 40.0% | 11 |
| Not Documented | 0 | 0.0% | 0 | 0.0% | 1 | 6.7% | 1 |
| Extent of Surgery |  |  |  |  |  |  |  |
| Gross-Total | 4 | 50.0% | 7 | 77.8% | 6 | 40.0% | 17 |
| Subtotal | 3 | 37.5% | 2 | 22.2% | 7 | 46.7% | 12 |
| Not Documented | 1 | 12.5% | 0 | 0.0% | 2 | 13.3% | 3 |
| Hemisphere |  |  |  |  |  |  |  |
| Left | 1 | 12.5% | 4 | 44.4% | 9 | 60.0% | 14 |
| Right | 6 | 75.0% | 5 | 55.6% | 4 | 26.7% | 15 |
| Not Documented | 1 | 12.5% | 0 | 0.0% | 2 | 13.3% | 3 |
| Primary Lobe |  |  |  |  |  |  |  |
| Frontal | 2 | 25.0% | 7 | 77.8% | 7 | 46.7% | 16 |
| Temporal | 2 | 25.0% | 1 | 11.1% | 6 | 40.0% | 9 |
| Parietal | 4 | 50.0% | 1 | 11.1% | 2 | 13.3% | 7 |

### Overall Survival

The mean number of nine-week cycles for each cohort was 4.6, 4.1, and 2.1 for vaccine, placebo, and ancillary groups, respectively. Overall survival (OS) at one year (OS-12) was the primary endpoint and was calculated based on time of post-surgical registration until death. No clinically significant differences were observed in OS trends, with those receiving placebo vs vaccine showing nearly overlapping survival curves, though interestingly both outperforming the ancillary group that in principle received the same pembrolizumab treatment and clinical follow-up as the placebo group (Figure 2A). Median OS was 14.4 months for the vaccine group, 15.2 months for the placebo group, and 8.8 months for the ancillary group (Figure 2B). OS-12 was 65.6% (35.2-100%) for the vaccine group and 75.0% (50.3-100%) for the placebo group (Figure 2C).

**Figure 2:**
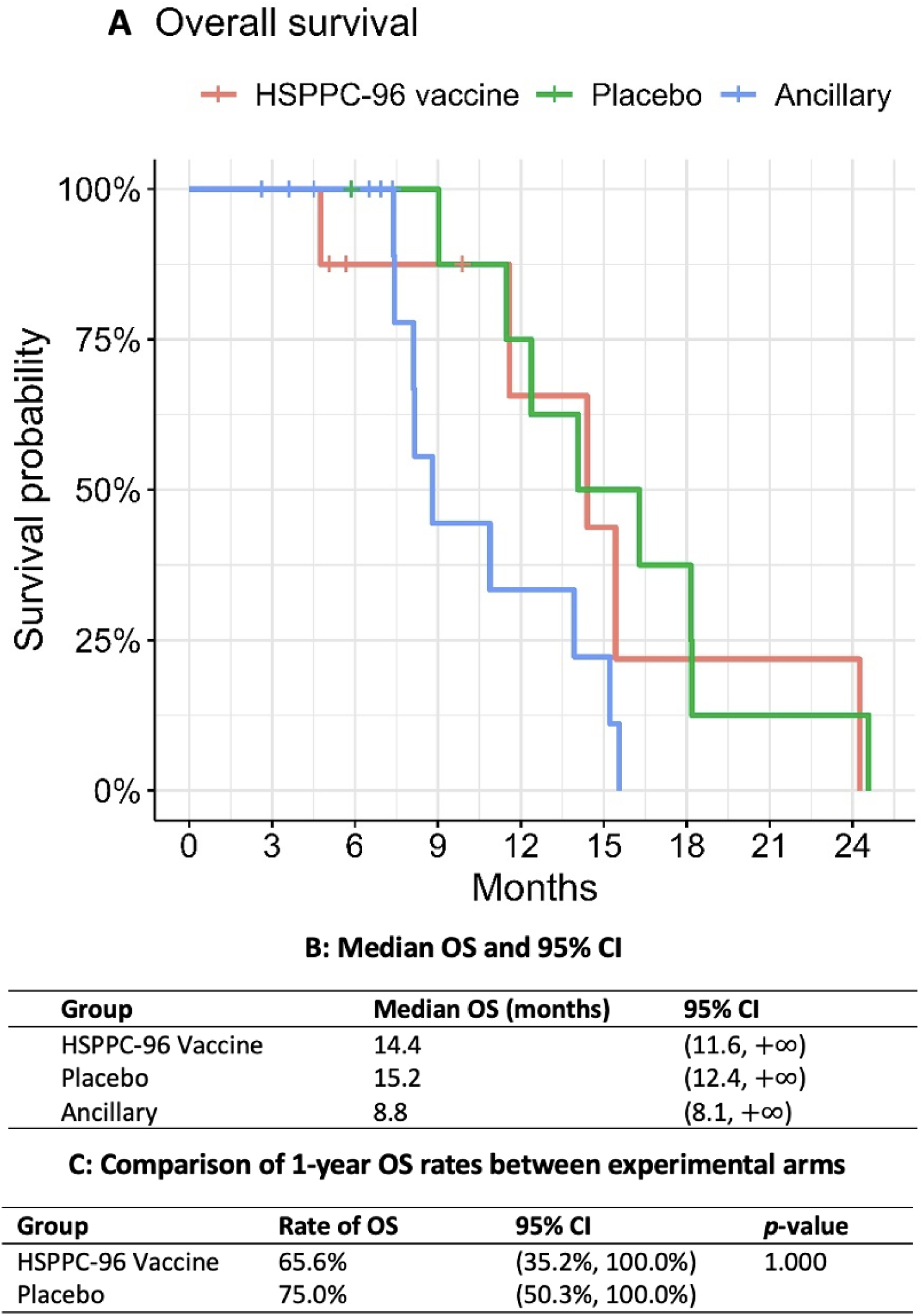
Kaplan-Meier plots comparing overall survival between the three treatment groups: HSPPC-96 Vaccine (red), Placebo Vaccine (green), and Ancillary Treatment of Pembrolizumab-only (blue) (A). Median overall survival times (B) and overall survival at 1 year (C) are listed below the Kaplan-Meier plots.

### Progression-Free Survival

Progression-free survival (PFS) and at six months (PFS-6) were both secondary endpoints in this clinical trial, calculated based on time of registration until disease progression or death. In contrast to OS plots, patients receiving HSPPC-96 vaccine demonstrated slightly prolonged PFS in vaccine groups compared to both placebo and ancillary groups (Figure 3A). Though not statistically significant and with overlapping 95% confidence intervals, the median PFS for the vaccine group was 9.9 months compared to nearly identical rates among the placebo group at 7.9 months and the ancillary group at 7.4 months (Figure 3B). The PFS-6 similarly shows a non-significant trend toward improvement, with the vaccine group at 87.5% (63.3-100%) compared to placebo at 55.6% (31.0-99.7%) and ancillary at 62.9% (41.6-95%) (Figure 3C).

**Figure 3:**
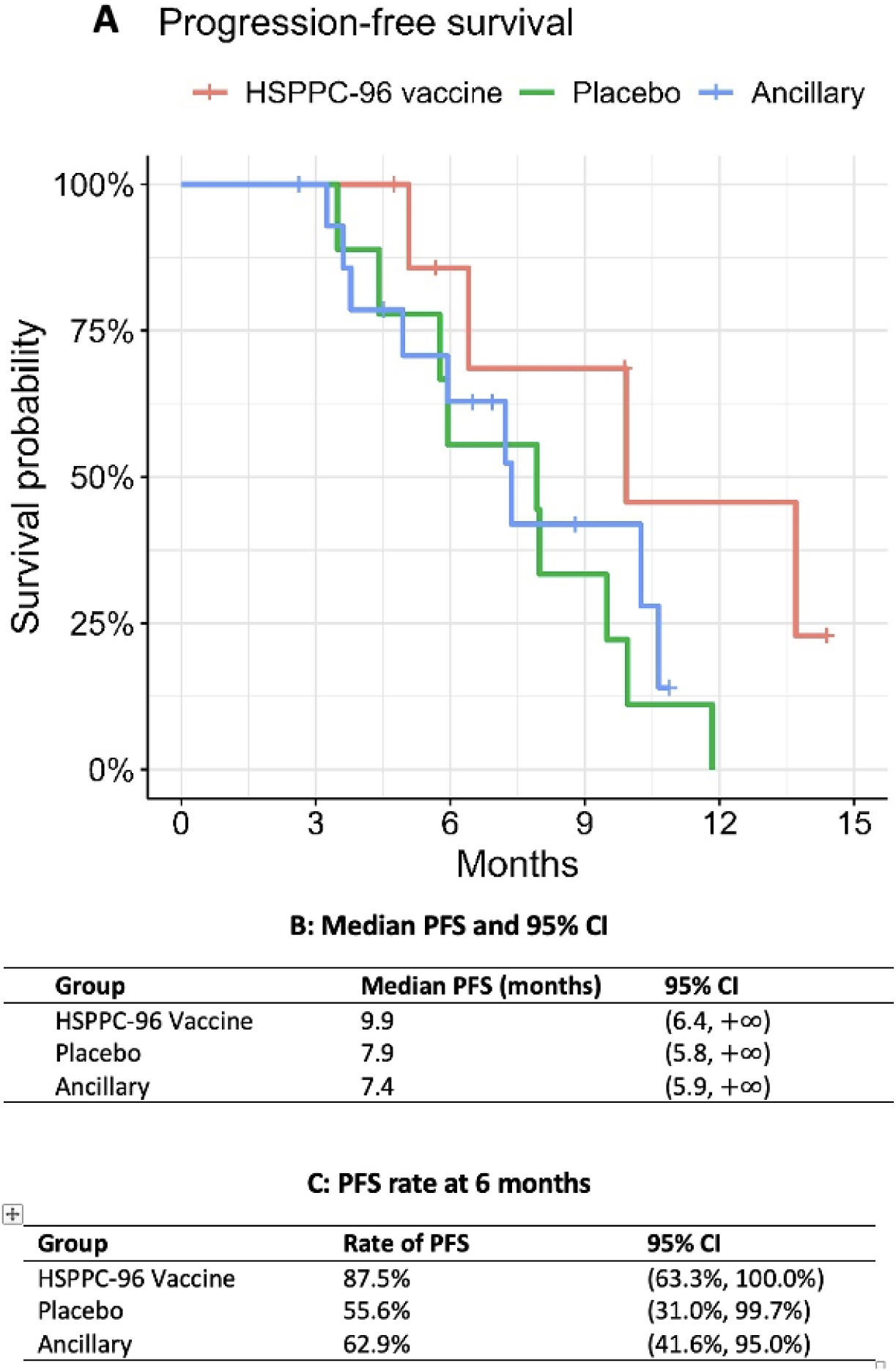
Kaplan-Meier plots comparing progression-free survival between the three treatment groups: HSPPC-96 Vaccine (red), Placebo Vaccine (green), and Ancillary Treatment of Pembrolizumab-only (blue) (A). Median progression-free survival times (B) and progression-free survival at 6 months (C) are listed below the Kaplan-Meier plots.

### Response Rates

Response rates were determined by RANO criteria for response^14^ (Table 2). Of the 32 total patients accrued, 26 were evaluable for radiographic responses. The majority across all groups had stable disease as their best responses. Within the experimental groups, stable disease was the best response in both groups, with six (100%) in the vaccine cohort and eight (88.9%) in the placebo group. One patient (11.1%) in the placebo group had progressive disease as their best response. The ancillary group had one partial response (9.1%) but also seven stable disease (63.6%) and three progressive disease (27.3%) responses.

**Table 2.** Radiographic Response Rates.

| Response rates and 95% CI in each category |  |  |  |  |
| --- | --- | --- | --- | --- |
| Group |  | Counts | Response rate | 95% CI |
| HSPPC-96 Vaccine | Total number of patients analyzed | 6 |  |  |
|  | Complete response | 0 | 0.0% | (0.0%, 45.9%) |
|  | Partial response | 0 | 0.0% | (0.0%, 45.9%) |
|  | Progressive disease | 0 | 0.0% | (0.0%, %) |
|  | Stable disease | 6 | 100.0% | (54.1%, 100.0%) |
| Placebo | Total number of patients analyzed | 9 |  |  |
|  | Complete response | 0 | 0.0% | (0.0%, 33.6%) |
|  | Partial response | 0 | 0.0% | (0.0%, 33.6%) |
|  | Progressive disease | 1 | 11.1% | (0.3%, 48.2%) |
|  | Stable disease | 8 | 88.9% | (51.8%, 99.7%) |
| Ancillary | Total number of patients analyzed | 11 |  |  |
|  | Complete response | 0 | 0.0% | (0.0%, 28.5%) |
|  | Partial response | 1 | 9.1% | (0.2%, 41.3%) |
|  | Progressive disease | 3 | 27.3% | (6.0%, 61.0%) |
|  | Stable disease | 7 | 63.6% | (30.8%, 89.1%) |

### Adverse Events

Twenty-one participants out of 32 accrued had reportable adverse events (AE) that were Grade 3 or 4, with 18 of these patients having reportable AEs attributable by the study team to at least one of the components of the study treatment (TMZ, Pembro, radiation therapy, vaccine, or surgery). Of these 18, four were in the vaccine group (50% of vaccine participants), five in the placebo group (56% of placebo participants), and nine in the ancillary group (60% of ancillary participants). Across all cohorts, lymphopenia was the prevailing grade 3 toxicity, attributed most frequently to temozolomide. Transaminitis and thrombocytopenia were the next most common grade 3/4 toxicity, with none in the vaccine group and most likely attributable to temozolomide. All grade 3/4 toxicities and percentages attributed to study treatment are listed in Table 3.

**Table 3.**
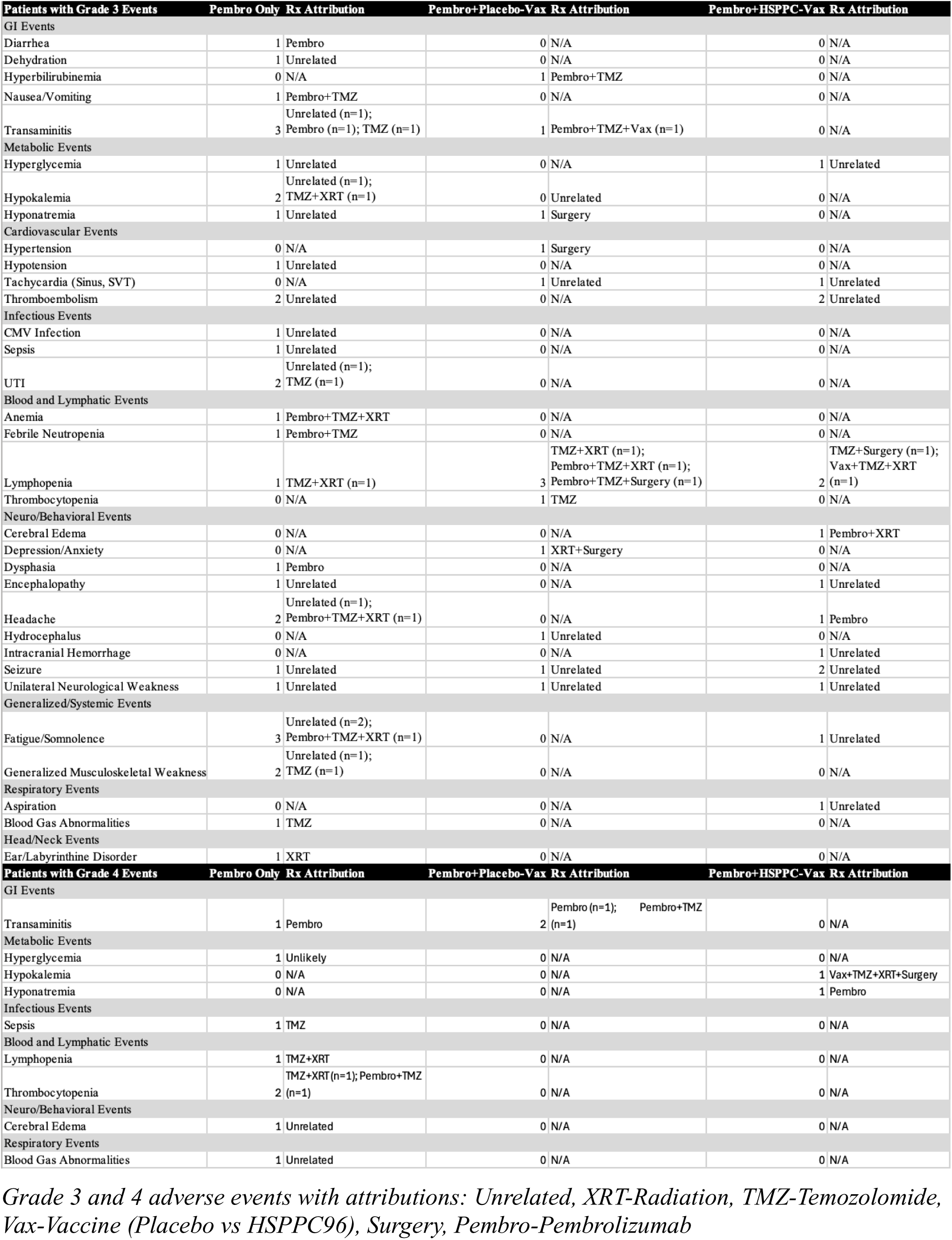

### Patient-Reported Outcomes

Of the 32 patients accrued to this protocol, only 30 completed baseline evaluations. Of these, a total of 13 patients (43%) provided complete survey responses from baseline through adjuvant cycle 2, the earliest timepoint to accurately assess patient-reported outcomes (Table 4). The self-reported symptom severity and interference with daily activities parameters from the MDASI-BT were evaluated to describe the magnitude and variability of symptom severity over the treatment course and between treatment arms. The results are underpowered due to small numbers of survey completion and study design, but the linear mixed models did not show any significant difference between treatment arms for the 10 outcome variables. However, there was a significant effect for cognitive, treatment-related, overall interference, and mood-related interference symptoms over time. The Bonferroni method indicated that baseline values of these parameters were significantly lower (i.e., less symptomatic) compared to timepoints prior to adjuvant cycle 1 (i.e., post-chemoradiation and immediately prior to adjuvant TMZ+Pembro with/without vaccine) as well as post adjuvant cycle 1 (Table 5). A sensitivity analysis using two different time points as end points (pre-Cycle 1 and post-Cycle 2) showed similar results (Supplemental Table 1).

**Table 4.** Patient-Reported Outcomes.

|  | Treatment Arm |  |  |  |
| --- | --- | --- | --- | --- |
| Time Point | Ancillary | Vaccine | Placebo | Total |
| Baseline/Prior to RT | 14 | 8 | 8 | 30 |
| Prior to adjuvant cycle 1 | 5 | 5 | 6 | 16 |
| Adjuvant post cycle 1 | 3 | 6 | 7 | 16 |
| Adjuvant post cycle 2 | 3 | 4 | 6 | 13 |
| Adjuvant post cycle 3 | 2 | 4 | 3 | 9 |

**Table 5.** Linear mixed models for symptom severity, symptom factor, and interference scores using Patient-Reported Outcomes surveys.

| Outcome | Treatment arm estimated mean |  |  | Treatment arm p-value | Timepoints estimated mean |  |  |  | Timepoints P-value | Treatment arm by timepoint interaction p-value |
| --- | --- | --- | --- | --- | --- | --- | --- | --- | --- | --- |
|  | Ancillary Mean | Vaccine Mean | Placebo Mean |  | Baseline Mean | Prior to adj cycle 1 Mean | Adj post cycle 1 Mean | Adj post cycle 2 Mean |  |  |
| Overall symptom burden | 1.53 | 2.13 | 1.63 | .459 | 1.26 | 1.88 | 2.12 | 1.78 | .064 | .883 |
| Affective | 1.76 | 3.09 | 2.35 | .124 | 1.99 | 2.40 | 2.79 | 2.41 | .535 | .739 |
| Cognitive | 1.98 | 2.38 | 1.66 | .670 | 1.21<sup>A</sup> | 2.19<sup>B</sup> | 2.62<sup>B</sup> | 2.0 | .034 | .331 |
| Neurologic | 1.24 | 1.48 | 1.27 | .908 | 1.28 | 1.31 | 1.47 | 1.26 | .926 | .874 |
| Treatment-related | 1.33 | 2.61 | 2.15 | .213 | 1.29<sup>A</sup> | 2.5<sup>B</sup> | 2.48<sup>B</sup> | 1.84 | .022 | .582 |
| General disease | 1.35 | 1.49 | 1.10 | .798 | .9 | 1.16 | 1.66 | 1.54 | .279 | .294 |
| GI | .123 | .97 | .59 | .361 | .21 | .59 | .70 | .75 | .515 | .556 |
| Overall Interference | 2.92 | 3.91 | 2.87 | .689 | 2.34<sup>A</sup> | 3.77<sup>B</sup> | 3.91<sup>B</sup> | 2.93 | .011 | .763 |
| Mood-related interference | 2.63 | 3.30 | 1.92 | .652 | 1.43<sup>A</sup> | 3.66<sup>B</sup> | 3.18<sup>B</sup> | 2.20 | .013 | .785 |
| Activity-related interference | 2.99 | 4.59 | 3.4 | .561 | 3.17 | 4.49 | 4.15 | 2.86 | .058 | .884 |

## Discussion

Glioblastoma is one of the most aggressive solid tumors across cancers and treatment advances are complicated not just by its molecular complexity and blood-brain barrier inaccessibility but by an immune microenvironment that is suppressive. The first major advance in the use of immunotherapy was the discovery of anti-PD1 immune checkpoint therapy, which demonstrated evidence of efficacy in solid tumors including with brain metastases^15,16^. Unfortunately, numerous Phase 2 and 3 studies in GBM failed to demonstrate efficacy in the upfront treatment setting when combined with chemoradiation^17^. Intriguing results from the neoadjuvant use of Pembro prior to surgery for recurrent GBM did demonstrate a modest benefit and suggested there may be potential for a tumor immune microenvironment to be modified by anti-PD1 therapy^18^, but more recent results from an expansion cohort did not reproduce the findings of improved clinical outcomes^19^. In toto, GBM has not benefited from checkpoint inhibitor immunotherapeutic treatment advances in the way other solid tumors have, and innovative approaches are needed to determine methods that may better capitalize on the GBM’s unique immune microenvironmental profile.

This Phase 2 study explored the use of the anti-PD1 agent Pembro administered during both the chemoradiation and adjuvant TMZ periods, with or without the addition of a novel personalized vaccination derived from the HSPPC-gp96 component of patient tumors. The primary endpoint of this study was overall survival at one year, with progression-free survival at 6 months, overall survival, progression-free survival, and radiographic response rates as secondary outcomes. Unfortunately, the study presented many logistical challenges in developing and administering the personalized vaccine component, especially as part of a multicenter effort in the setting of the pandemic, and accrual was closed prior to reaching participation numbers necessary to evaluate the protocol outcomes with appropriate power. Nonetheless, despite the low accrual rates, we failed to detect meaningful differences in the primary outcome OS-12 as well as in median OS among the treatment groups, and both treatment and placebo arms were within statistical margins from the Pembro-only ancillary group. PFS-6 (87.5% vs 55.6% and 62.9%) and median PFS (9.9 vs 7.9 and 7.7 months) were improved in the vaccine group compared to the placebo and ancillary groups, respectively, suggesting the possibility of benefit in the vaccine group. Furthermore, the stable disease rate of 100% in the vaccine group outperformed the other treatment arms. However, we encourage to interpret these findings with caution because of both the small sample sizes in each arm as well as the lack of demonstrable benefit when considering the more clinically robust outcome of OS. Even with new imaging interpretation criteria accounting for immunotherapies, PFS and response rates are notoriously difficult to interpret due to known limitations in MR imaging techniques in differentiating inflammation associated with recovery from radiation therapy or responses to immunotherapy from true tumor progression^20,21^.

By comparison, a recent publication using an alternative anti-PD1 checkpoint inhibitor Nivolumab as part of the CheckMate 498 study^22^ found similar clinical outcomes in their group of upfront nivolumab plus radiation-therapy group, with median OS at 14.9 months and median PFS at 6.2 months, similar to the 14.4-15.2 months and slightly shorter than the 7.7-9.2 months reported in our pooled study patients, respectively. This is in keeping with evolving literature disputing the efficacy of PD1-directed immunotherapies as monotherapy in the setting of GBM that drove our hypothesis to combine with other immune-activating agents. Unfortunately, it does not appear that HSPPC-96 vaccination combined with Pembro-based immunomodulation in this study sufficiently altered the immune landscape nor enhanced the survival outcomes or response rates.

We acknowledge that the early closure of this study due to recruitment and logistical difficulties resulted in too little statistical power to be able to state with any certainty whether this treatment was likely to show benefit or is truly ineffective. Simultaneously, we acknowledge that the lack of even isolated cases of benefit in the vaccine-treatment group is discouraging. Furthermore, since the inception of this study, additional literature has continued to demonstrate the limited efficacy of PD1-directed therapies in GBM. We are therefore unenthusiastic about continuing this line of inquiry using these agents in combination.

However, we do not endorse abandonment of the HSPPC-96 or other personalized vaccination strategies writ large. For one, the use of the vaccine appears to be safe. No significant differences in adverse events and toxicities were observed between the treatment arms, with rates of Grade 3/4 toxicity similar between arms and none clearly attributable to vaccination, suggesting the administration of the novel compound was safe. Similarly, the Grade 3/4 toxicities attributable to any component of the therapy (e.g., radiation, TMZ, Pembro, or vaccine) across all groups were ones common to TMZ and/or radiation administration, suggesting that there was no clear increase in major adverse events with the addition of Pembro with or without vaccine during the study period. Secondly, the strategy of individualized antigen-presentation and immune activation through vaccination engagement of APCs^23^ or direct engineering of APCs like dendritic cells^24^ remains a compelling hypothesis^25^. The combination of this vaccine with Pembro does not appear to be effective, and this may in part be due to both highly variable expression rates of PDL1 in GBM as well as lack of demonstrable correlation to OS^26^, suggesting PD1/PDL1-axis inhibition may be the wrong combinatorial strategy. Instead emerging targets such as B7-H3 may be a more attractive co-target; it is highly expressed in GBM and its microenvironment^27,28^, contributes to tumorigenesis by both immune and non-immunological mechanisms^29^, and also has blocking antibodies^30^, antibody-drug conjugates, and CAR-T strategies already in development^29^. Targets such as B7-H3 and others may benefit from antigen-presentation enhancing strategies such as HSPPC vaccine described here and should be considered as part of the ongoing pipeline of combinatorial immunotherapeutic interventions in GBM.

In conclusion, this trial combining upfront chemoradiation therapy with Pembro followed by adjuvant TMZ and Pembro with or without HSPPC vaccine failed to meet its primary endpoint of OS-12 and only modest improvements in PFS. The regimen overall was well-tolerated but presented significant logistical difficulties in the production and administration of vaccine. Although we cannot advocate for revamping this trial in this combination, we nonetheless encourage ongoing exploration of personalized vaccines with other rationally chosen immunotherapeutic interventions to continue exploring mechanisms of further unlocking the potential for immunotherapy in GBM and other CNS tumors.

## Authorship

Results were collated and manuscript was written by B.H.O. and provided to all authors for review and to provide revisions. Trial participants were enrolled, trial duties were executed, and data was collated, analyzed, interpreted, and reported as part of primary and/or sub-investigator obligations by S.M.L., R.T.M., C.R.T., J.D.R., N.G.A., N.R., S.K., Y.R., T.W., E.P., M.Y., K.L.F., J.J.M., L.P.T., H.C., E.M.D., N.P., M.R.G. Original trial design was by J.J.R. Drug supply/manufacture came from E.V. and S.C. from Merck and Agenus, respectively. Biostatistical support and analyses were by Y.Y. Multicenter regulatory and nursing coordination provided by C.C. and K.W., respectively. Central data management provided by E.G. Outcomes data analysis performed by T.M. and T.S.A. Central associate investigator duties and trial accrual and management support was provided E.C.B., H.E.L., J.G., and M.P.-P.

## Data Availability

the data will be made available upon reasonable request to the corresponding author or designee.

## Acknowledgments

the authors would like to thank all research participants and the family and friends who supported them through the trial and the disease process. Many past and present researchers, clinicians, and staff have worked at each institution associated with the NCI Brain Tumor Trials Collaborative (BTTC) than can be highlighted and we thank all those who have been part of its research and mission. The opinions expressed in this paper are those of the authors and do not necessarily represent those of Merck Sharpe & Dohme Corp. Furthermore, the contributions of the NIH author(s) were made as part of their official duties as NIH federal employees, are in compliance with agency policy requirements, and are considered Works of the United States Government. However, the findings and conclusions presented in this paper are those of the author(s) and do not necessarily reflect the views of the NIH or the U.S. Department of Health and Human Services.

