## Supplemental Table for "Pembrolizumab, Temozolomide and HSPPC-96 Vaccine in Newly Diagnosed Glioblastoma Post-Chemoradiation: Results from a Multi-institutional, Phase 2, Randomized, Placebo-Controlled Trial"

Supplementary Table 1

| Outcome | Up to post adjuvant cycle 1 | | | Up to post adjuvant cycle 3 | | |
| --- | --- | --- | --- | --- | --- | --- |
|  | Treatment arm p-value | Timepoints P-value | Treatment arm by timepoint interaction p-value | Treatment arm p-value | Timepoints P-value | Treatment arm by timepoint interaction p-value |
| Overall symptom burden | .624 | .021 | .959 | .400 | .061 | .861 |
| Affective | .428 | .352 | .987 | .110 | .590 | .822 |
| Cognitive | .945 | .009 | .489 | .287 | .027 | .180 |
| Neurologic | .920 | .889 | .997 | .894 | .725 | .892 |
| Treatment-related | .127 | .008 | .376 | .318 | .031 | .710 |
| General disease | .933 | .159 | .389 | .481 | .323 | .258 |
| GI | .159 | .575 | .327 | .364 | .260 | .689 |
| Overall Interference | .790 | .007 | .669 | .613 | .011 | .693 |
| Mood-related interference | .739 | .006 | .532 | .464 | .013 | .720 |
| Activity-related interference | .704 | .095 | .042 | .433 | .055 | .852 |
